# Topographic Heterogeneity of Lung Microbiota in End-Stage Idiopathic Pulmonary Fibrosis: The Microbiome in Lung Explants-2 (MiLEs-2) Study

**DOI:** 10.1101/2020.03.05.20031021

**Authors:** Eleanor B Valenzi, Haopu Yang, John C Sembrat, Libing Yang, Spencer Winters, Rachel Nettles, Daniel J Kass, Shulin Qin, Xiaohong Wang, Michael M Myerburg, Barbara Methé, Adam Fitch, Jonathan K Alder, Panayiotis V Benos, Bryan J McVerry, Mauricio Rojas, Alison Morris, Georgios D Kitsios

## Abstract

**Background:** Lung microbiota profiles in patients with early idiopathic pulmonary fibrosis (IPF) have been associated with disease progression; however, the topographic heterogeneity of lung microbiota and their roles in advanced IPF are unknown.

**Methods:** We sampled subpleural tissue from up to three lobes as well as airway-based specimens (bronchial washings and airway tissue) in patients with IPF, connective tissue disease-associated interstitial lung disease (CTD-ILD), cystic fibrosis (CF), and chronic obstructive pulmonary disease (COPD) and donor lungs deemed unsuitable for transplant (controls). We quantified bacterial load and profiled communities by polymerase chain reaction (PCR) amplification and sequencing of the 16S rRNA gene.

**Findings:** Explants from 62 IPF, 15 CTD-ILD, 20 CF, 20 COPD and 20 control patients were included. Airway-based samples had higher bacterial load compared to distal parenchymal tissue across all patient groups. IPF basilar tissue had much lower bacterial load compared to CF and control lungs (p<0.001). Among patients with IPF, no differences in microbial community profiles were found between parenchymal tissue samples from different lobes. With Dirichlet multinomial models, a cluster of IPF patients (29%) with distinct composition, high bacterial load and low alpha diversity was identified, exhibiting higher odds for acute exacerbation of IPF or death.

**Interpretation:** IPF explants exhibited low biomass in the distal parenchyma of all three lobes with higher bacterial load in the airways. The discovery of a distinct subgroup of IPF patients with higher bacterial load and worse clinical outcomes supports investigation of personalized medicine approaches for microbiome-targeted interventions.

**Key Messages:**

- What is the key question? Bronchoalveolar lavage microbiome profiles in early idiopathic pulmonary fibrosis (IPF) have been associated with disease progression, but the regional heterogeneity of resident microbiota in end-stage IPF has not been defined.
- What is the bottom line? IPF explants demonstrate higher bacterial load in airway compared to parenchymal samples, but no differences in between apical or basilar parenchymal samples. A subgroup of patients with higher bacterial load and respiratory pathogen abundance was associated with worse clinical outcomes.
- Why read on? Patient-specific heterogeneity in the lung microbiome of IPF supports the need for personalized microbiome-targeted interventions in IPF.

## INTRODUCTION

Idiopathic pulmonary fibrosis (IPF) is a devastating age-associated disease, occurring more frequently in smokers and carriers of host-defense gene mutations.[1, 2] While it is theorized that alveolar injury in a genetically-susceptible host propagates aberrant repair mechanisms resulting in fibrosis, the exact environmental factors provoking lung injury have not been defined.[3] Dysbiosis in the respiratory tract has been proposed as a potential mechanism for precipitation and/or perpetuation of lung injury. The hypothesis that lung microbiota contributes to IPF progression emerged from epidemiologic observations suggestive of host-microbiome interactions in the respiratory tract, such as the finding that immunosuppression increases mortality in IPF, whereas antibiotics may offer survival benefit in treatment-tolerant and adherent IPF patients.[4-6] Three prospective cohort studies in patients with early IPF provided direct evidence for the lung microbiome hypothesis with the use of culture-independent, bacterial DNA sequencing techniques. Bronchoalveolar lavage (BAL) fluid from patients with IPF had higher bacterial burden compared to chronic obstructive pulmonary disease (COPD) patients and healthy controls,[7] and those IPF patients with the highest bacterial burden exhibited worse outcomes.[7-9] Lung microbial profiles were associated with distinct host transcriptome responses.[10, 11] Furthermore, in murine model of fibrosis, lung dysbiosis preceded peak lung injury and was associated with worse survival.[8] Therefore, lung microbiota manipulation with antibiotic therapies has become an attractive target for intervention, currently assessed by ongoing clinical trials.[12, 13]

The role of lung microbiota in later stages of IPF remains unknown. Furthermore, the involvement of microbes across the respiratory tract in such a disease hallmarked by spatial heterogeneity (subpleural predominance and an apicobasal gradient of fibrosis) is poorly understood. The BAL samples used by prior studies capture microbiota from lower generations of the tracheobronchial tree and up to 5% of the alveolar space, but do not provide detailed regional characterization of microbiota. In a previous study from our group (Microbiome in Lung Explants - MiLEs study), we examined distal parenchymal tissue from lung explants of patients with end-stage usual interstitial pneumonia (UIP) at the time of lung transplant or death.[14] Surprisingly, we found exceedingly low bacterial signals by 16S rRNA gene sequencing, in contrast to explant tissue from cystic fibrosis (CF) or donor lungs. These findings suggest that advanced honeycombing may represent not only a physiologic dead-space, but also an ecological dead-space (“*microbiome desert*”), whereas microbiota may primarily colonize the airways and areas of traction bronchiectasis.[14] The discordance between IPF bacterial burden in BAL versus in distal parenchymal tissue suggests that fibrosis and the resulting honeycombing could be a maladaptive fibrotic response to airway microbiota.[15]

To gain further understanding of the spatial heterogeneity of lung microbiota in IPF, our current study (MiLEs-2) characterized the regional tissue microbiome across the apicobasal axis of UIP as well as the airways in lung explants from patients with IPF. In particular, we sampled up to three different lobes and airway-based samples in IPF explants, as well as similar samples from explants with connective tissue disease-associated interstitial lung disease (CTD-ILD), CF, COPD, and lungs that had been donated but rejected for transplant (control samples).

## METHODS

### Study design

MiLEs-2 is a retrospective, case-control study of explanted lung tissue obtained at the time of lung transplantation or rapid autopsy from patients with IPF and diseased controls (CTD-ILD, CF, and COPD). We also obtained control tissue samples from lung donation candidates deemed unsuitable for transplant, via the Center for Organ Recovery and Education (CORE). IPF diagnoses were confirmed according to 2018 clinical practice guidelines, with histopathology results from lung explants (and prior surgical lung biopsies for some cases) reviewed by specialized thoracic pathologists at the University of Pittsburgh Medical Center.[16] We classified patients with IPF as those with an acute exacerbation of IPF (AE-IPF) versus chronic IPF per established criteria.[14, 17] Informed consents for conducting research utilizing the explanted lung specimens were obtained from patients or their designated representatives. The University of Pittsburgh Institutional Review Board and Committee for Oversight of Research and Clinical Training Involving Decedents approved this study.

### Sample acquisition and processing

We obtained lung tissue and airway samples in the operating room or autopsy suite per established protocols.[14] We resected a subpleural lower lobe tissue segment, which was further dissected into pieces weighing an average of 45mg under sterile conditions (Figure 1). For a random subset of explants based on logistical feasibility, we also resected subpleural tissue from the right middle lobe or lingula (for right or left lung explants respectively) and the upper lobe. For a smaller subset of explants, we also collected a bronchial wash specimen (by aspiration of 30mL of phosphate-buffered saline instilled into a bronchial segment using a sterile tube) as well as an airway tissue specimen (from a segmental bronchus) prior to parenchymal tissue sample collection. Samples were frozen in liquid nitrogen and stored at −80°C until processing.

**Figure 1.**
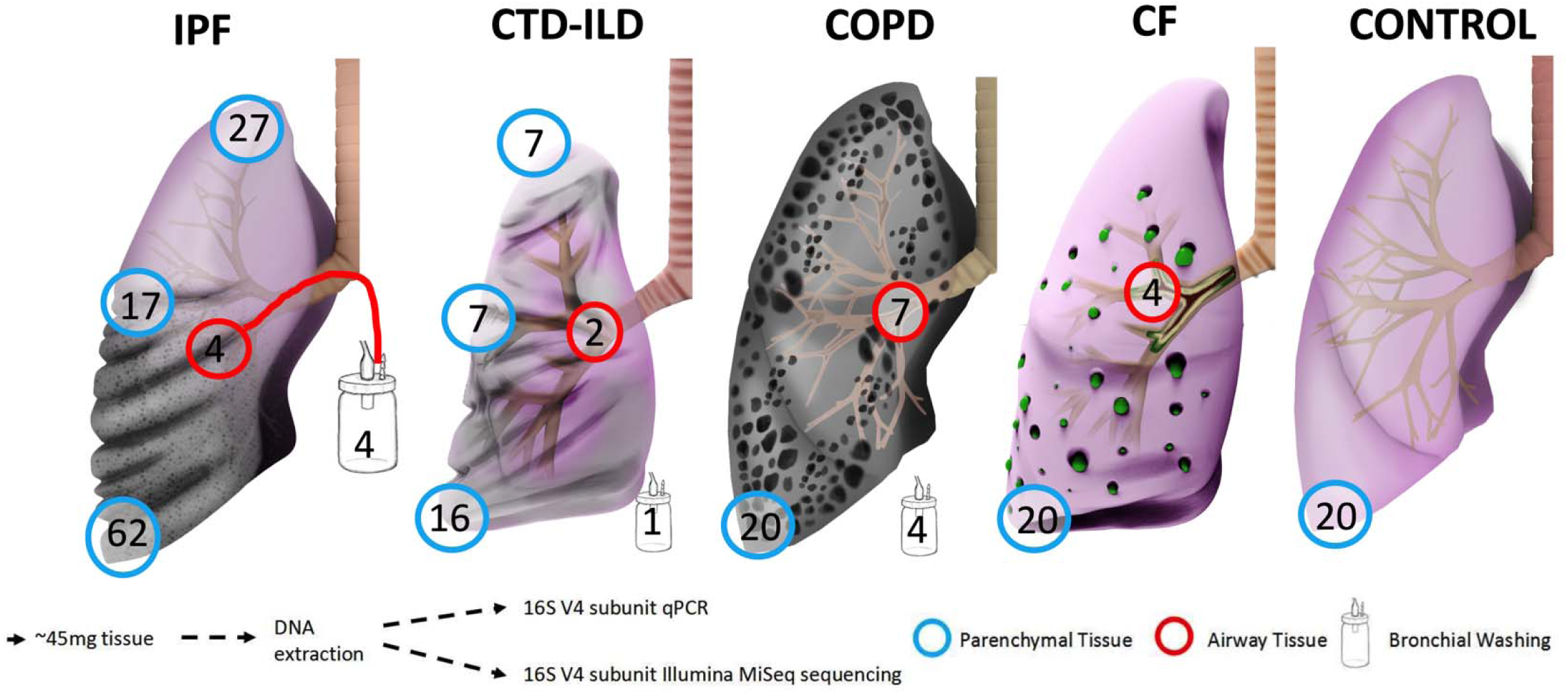
Explanted lung tissue and bronchial washing samples included in the study, depicted by disease and tissue anatomic location. Numbers in the circles indicate available number of samples from each anatomic site or type. IPF, idiopathic pulmonary fibrosis; CTD-ILD, connective tissue disease-associated interstitial lung disease; COPD, chronic obstructive pulmonary disease; CF, cystic fibrosis.

### DNA extraction, 16S rRNA qPCR and pyrosequencing

We extracted genomic DNA and performed polymerase chain reaction (PCR) amplification of the V4 hypervariable region of the 16S rRNA gene.[18] Amplicons of the V4 rRNA bacterial gene subunit were sequenced on the Illumina MiSeq platform and quantified by qPCR.[14] We also quantified the human genomic DNA present in each sample by qPCR of the human Glyceraldehyde 3-phosphate dehydrogenase (*GAPDH*) gene. We conducted a series of experiments with lung explant tissue to identify optimal sample type (whole tissue vs. swab) and ruled out the presence of PCR inhibitors (Supplement).

### Statistical analysis

From derived 16S sequences, we applied a custom pipeline for Operational Taxonomic Units (OTUs-taxa) classification (Supplement) and performed analyses at genus level. We calculated descriptive statistics of clinical characteristics and performed nonparametric comparisons using the R software (v.3.5.1). Ecological analyses of alpha diversity (Shannon index) and beta diversity (Bray-Curtis index with permutational analysis of variance [permanova] at 1000 permutations) were conducted using the R *vegan* package and visualized with principal coordinates analyses (PCoA) plots. For comparisons with lower respiratory communities in healthy controls, we utilized 16S sequencing data from a previous study that had analyzed BAL specimens from healthy volunteers.[19] For 30 IPF patients who had survived to lung transplantation and had whole genome sequencing performed in genomic DNA extracted from blood samples, we obtained genotypes for the promoter single nucleotide polymorphism (SNP) rs35705950 of the *MUC5B* gene. To agnostically examine for distinct clusters of microbial composition (“meta-communities”) in our IPF basilar tissue samples (n=62), we applied unsupervised Dirichlet Multinomial Models (DMM) with Laplace approximations to define the optimal number of clusters in our dataset.[20] We then examined for associations of microbiome variables (bacterial load [log-transformed end-point fluorescence of qPCR assay], alpha diversity, beta-diversity and DMM clusters) with clinical variables (disease classification, diagnosis of AE-IPF, lung transplant vs. death outcome and *MUC5B* genotypes).

## RESULTS

### Study population

We analyzed basilar lung explant tissue specimens from 62 IPF patients, 15 CTD-ILD patients, 20 CF patients, 20 COPD patients and 20 control lungs (CORE). Additional specimens (middle/upper lobe tissue or airway-based samples) were available from a subset of explants (Figure 1). Comparisons of clinical characteristics showed that a higher percentage of IPF patients were male (81%) (Table 1), and that IPF patients had severely decreased forced vital capacity (FVC) (median 41%) and diffusing capacity for carbon monoxide (DLCO) (median 30%), reflecting their end-stage status at the time of lung transplantation (76%) or rapid autopsy (24%). AE-IPF was clinically diagnosed in 35% of patients with IPF and was strongly associated with the finding of diffuse alveolar damage on explant histopathology (odds ratio [OR] = 9.8, 95% confidence interval [CI]: 2.5-44.3, p<0.0001).

**Table 1.** Clinical data for patients included in the MiLEs-2 cohort. Data presented as medians (Interquartile range [IQR]) or N (%).

| Variable | IPF | CF | COPD | CTD-ILD |
| --- | --- | --- | --- | --- |
| **N** | 62 | 20 | 20 | 15 |
| **Age (years), median (IQR)** | 64.5 [60.0, 69.0] | 29.0 [23.8, 35.0] | 62.0 [59.0, 65.0] | 63.0 [54.0, 68.5] |
| **Male, n (%)** | 50 (81%) | 7 (35%) | 9 (45%) | 7 (47%) |
| **Ever-smokers, n (%)** | 42 (68%) | 0 (0%) | 20 (100%) | 10 (67%) |
| **Total pack-years, median (IQR)** | 10.2 [0.0, 30.0] | 0.0 [0.0, 0.0] | 57.5 [40.0, 65.5] | 10.0 [0.0, 26.8] |
| **FEV1 (L), median (IQR)** | 1.5 [1.2, 2.0] | 0.7 [0.6, 0.9] | 0.6 [0.5, 0.7] | 1.4 [1.2, 1.6] |
| **FEV1 predicted %, median (IQR)** | 53.0 [43.0, 67.0] | 22.0 [18.8, 24.2] | 25.0 [22.0, 30.2] | 52.0 [45.5, 68.0] |
| **FVC (L), median (IQR)** | 1.8 [1.4, 2.5] | 1.4 [1.1, 1.8] | 2.0 [1.8, 2.3] | 1.7 [1.5, 2.1] |
| **FVC predicted %, median (IQR)** | 41.0 [37.0, 58.0] | 32.5 [28.0, 41.2] | 58.5 [48.0, 66.0] | 47.0 [39.2, 60.2] |
| **DLCO (ml), median (IQR)** | 6.4 [4.7, 9.3] | 12.8 [11.8, 14.6] | 6.8 [5.1, 9.9] | 5.1 [4.6, 7.5] |
| **DLCO predicted %, median (IQR)** | 30.0 [22.0, 36.0] | 58.0 [49.0, 67.0] | 32.5 [26.8, 51.0] | 25.0 [20.5, 28.5] |
| **Mean PAP (mmHg), median (IQR)** | 24.0 [20.0, 28.0] | 25.0 [21.0, 33.0] | 21.5 [19.0, 27.0] | 24.5 [21.0, 38.8] |
| **GERD<sup>#</sup>, n (%)** | 48 (77%) | 19 (95%) | 12 (60%) | 15 (100%) |
| **Systemic steroids, n (%)** | 15 (24%) | 9 (45%) | 6 (30%) | 9 (60%) |
| **Inhaled corticosteroids, n (%)** | 11 (18%) | 8 (40%) | 19 (95%) | 5 (33%) |
| **Specific therapies:** |  |  |  |  |
| **Pirfenidone, n (%)** | 10 (16%) | 0 (0%) | 0 (0%) | 1 (7%) |
| **Nintedanib, n (%)** | 9 (15%) | 0 (0%) | 0 (0%) | 0 (0%) |
| **Other immunomodulator, n (%)** | 15 (24%) | 0 (0%) | 1 (5.0%) | 9 (60%) |
| **BAL + culture or recipient tissue culture\*, n (%)** | 7 (12%)<sup>§</sup> | 20 (100%) | 7 (35%) | 2 (13%)<sup>§</sup> |
| **Diffuse alveolar damage on explant pathology, n (%)** | 21 (36%)<sup>§</sup> | 0 (0%) | 0 (0%) | 5 (33%) |
| **Antibiotics in past 3 months, n (%)** | 25 (40%) | 20 (100%) | 9 (45%) | 9 (60%) |
| **Lung transplant, n (%)** | 47 (76%) | 20 (100%) | 20 (100%) | 10 (67%) |

<sup>#</sup> GERD definition: based on clinical history, available data from esophagogram or upper endoscopy, or prescription for proton pump inhibitor or histamine receptor 2 blocker.
\* All patients who underwent lung transplantation had a tissue culture of their resected main stem bronchial tissue, as per the institutional transplant protocol.
<sup>§</sup> Percentage calculated with denominator of all patients with available data (patients with unavailable data for each variable were removed from calculations).
Abbreviations: FEV1: forced expiratory volume in 1 second; FVC: forced vital capacity, 6MWD: 6-minute walk distance; DLCO: diffusion capacity of the lungs for carbon monoxide; GERD: gastroesophageal reflux disease; PAP: pulmonary artery pressure.

### Airway-based samples have higher bacterial load than corresponding parenchymal tissue

By 16S qPCR across all available samples, bronchial washings had higher bacterial load than corresponding airway tissue, which in turn had higher bacterial load than their counterparts in basilar parenchymal tissue (Figure 2A). This bacterial load gradient from the airways to the distal parenchyma was present within both IPF and COPD lungs (Figure S1). As for the amount of human DNA available in each sample (quantified by *GAPDH* qPCR), a reverse gradient was observed compared to bacterial DNA load, with tissue samples having much higher human DNA content compared to bronchial washings (p<0.0001) (Figure S2). Sample type was further associated with significant difference in alpha diversity (Shannon index, Figure 2B), with airway tissue samples having the lowest alpha diversity compared to the other types of samples, as well as significantly different taxonomic composition by beta-diversity comparisons (Figure 2C). Overall, airway-based samples captured higher microbial biomass with lower human DNA abundance, whereas distal parenchymal tissue (mainly from IPF and COPD explants) had lower biomass and higher amount of human DNA present.

**Figure 2.**
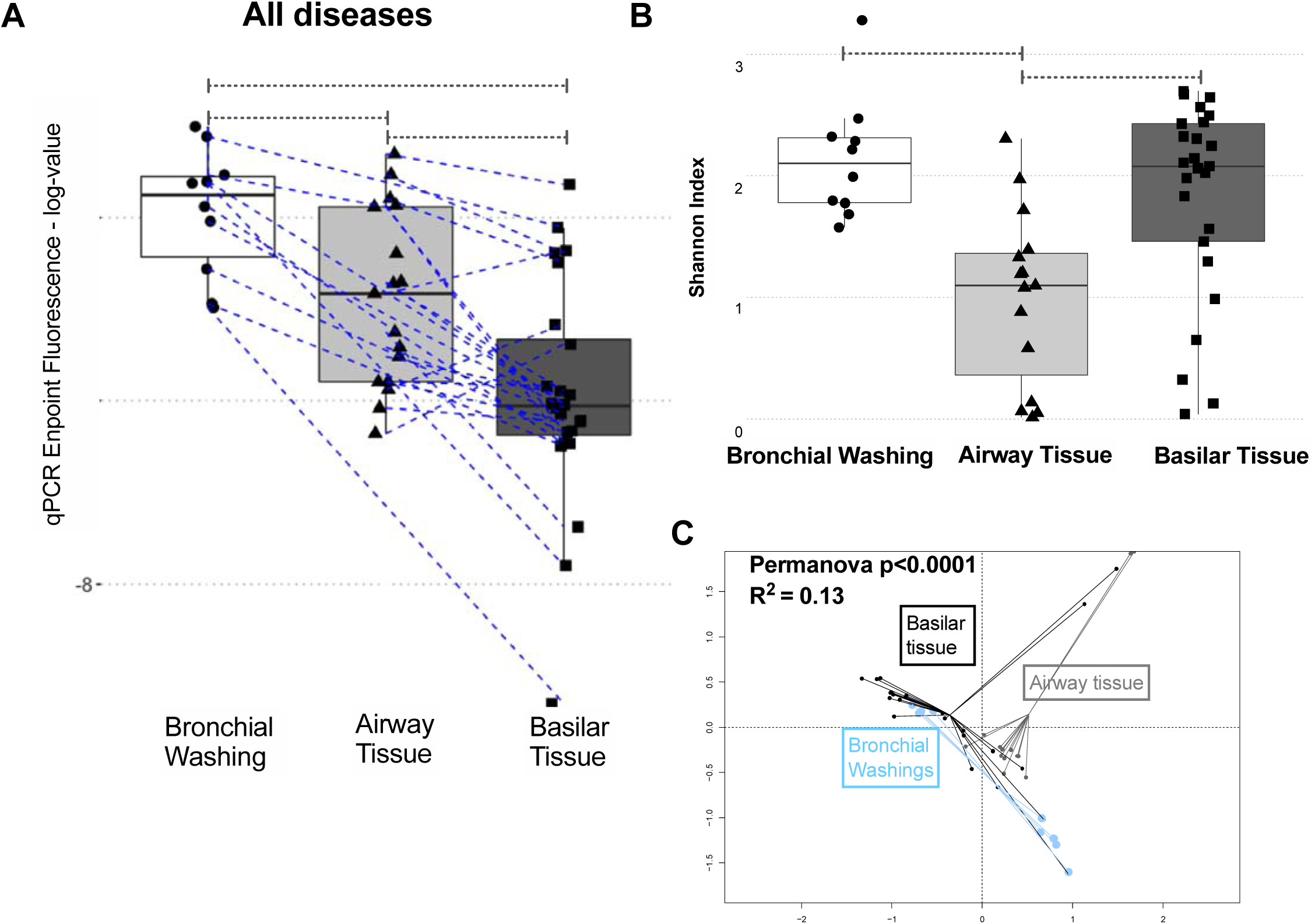
Airway-based samples have higher bacterial load compared to parenchymal tissue samples. (A) Bacterial load by qPCR endpoint fluorescence (log-transformed) in bronchial washings, airway tissue and basilar parenchyma tissue samples for all diseased samples available (IPF, CTD-ILD, COPD and CF). (B) Airway tissue samples had the lowest alpha diversity (Shannon Index) compared to bronchial washings or basilar tissue samples. (C) Significant compositional differences by beta diversity (Bray-Curtis dissimilarity index) comparisons visualized with Principal Coordinates Analysis plot between bronchial washings, airway tissue, and basilar parenchyma tissue samples. Pairwise p-values obtained from Wilcoxon tests. *= p<0.05, ***= p<0.001, ****= p<0.0001.

### IPF basilar tissue samples have low bacterial load

After demonstration of the airway-parenchyma gradient of bacterial load, we then examined for differences in bacterial load and community profiles in basilar tissue parenchymal specimens across the different disease states. By qPCR, IPF and COPD basilar tissue samples had much lower bacterial load (approximately 40-fold less bacterial DNA) compared to basilar parenchymal tissue from CF patients or CORE lungs (p<0.0001, Figure 3A). Notably, parenchymal samples in IPF and COPD have extensive anatomic destruction and physiologic dead-spaces (due to advanced honeycombing and emphysema, respectively), suggesting the presence of “ecological dead-spaces” for respiratory microbiota. These observations agree with our prior ones in a smaller cohort of end-stage IPF explants.[14] Examination of alpha diversity provided a reciprocal image of bacterial load: IPF samples had much higher Shannon index compared to CF samples (p<0.0001, Figure 3B). The pattern of low bacterial burden with high alpha diversity in IPF samples is strongly suggestive of experimental contamination due to low signal/noise ratio.[21] Significant taxonomic composition differences by disease were detected with beta-diversity comparisons (Figure 3C).

**Figure 3.**
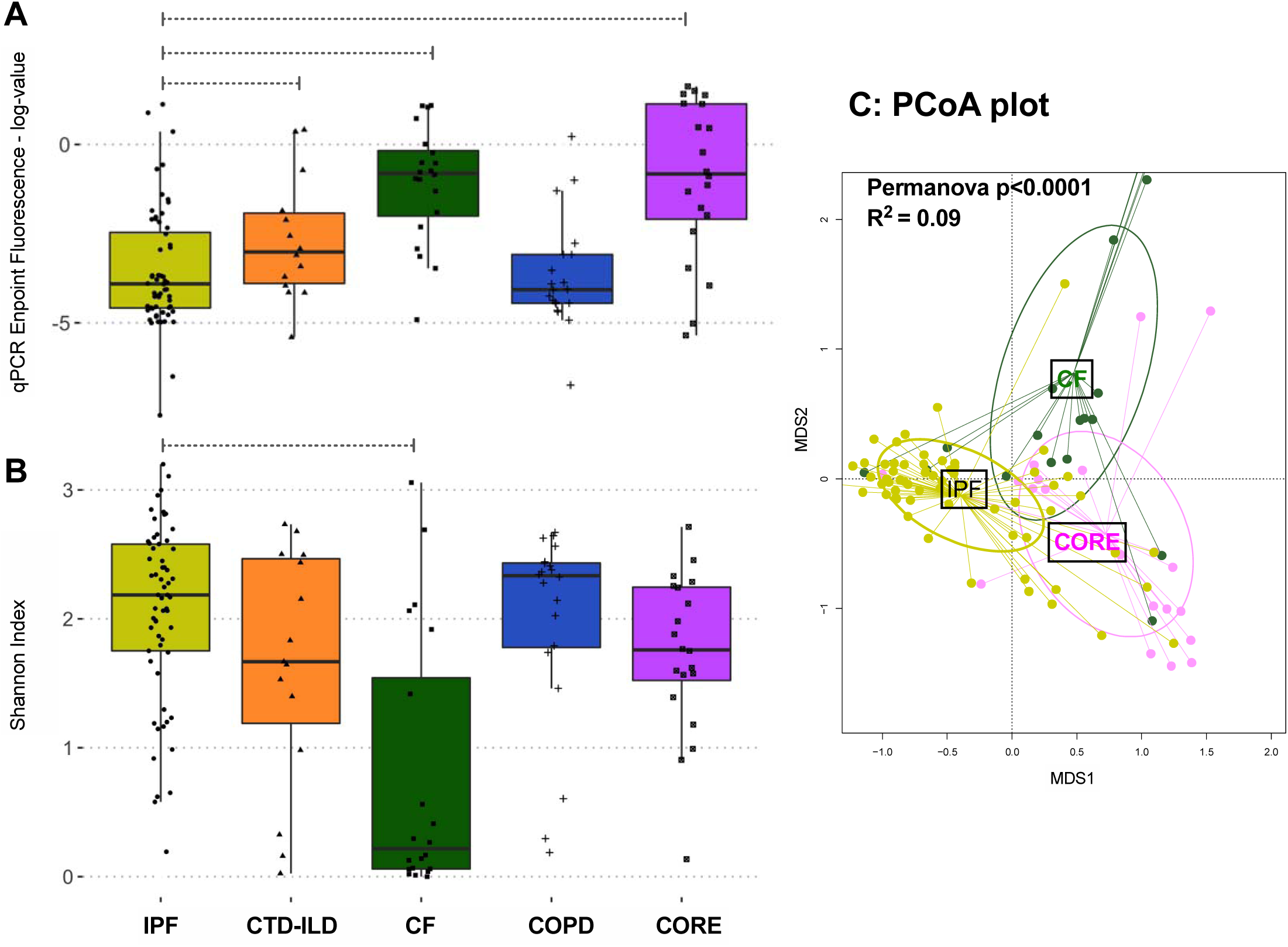
(A) Bacterial burden by qPCR endpoint fluorescence in basilar parenchyma tissue across all diseases and controls. (B) Alpha diversity, as measured by Shannon index, of basilar parenchyma tissue across all diseases and controls. (C) Beta diversity, as measured by Bray-Curtis dissimilarity in basilar parenchyma tissue of IPF, CF, and control lungs. P-values are obtained by Wilcoxon tests. *= p<0.05, ***=p<0.001, ****=p<0.0001. IPF, idiopathic pulmonary fibrosis; CTD-ILD, connective tissue disease-associated interstitial lung disease; COPD, chronic obstructive pulmonary disease; CF, cystic fibrosis; MDS1, primary NMDS axis; MDS2, secondary NMDS axis; NMDS, non-metric multidimensional scaling.

To further interpret the low bacterial signal from IPF basilar tissue samples, we examined the specific taxonomic composition of all samples available (Figure S3). We first focused on CF samples, which consisted of low diversity communities with a high abundance of typical pathogenic taxa (e.g. *Pseudomonas* or *Burkholderia* genera). Among 21 samples from CF patients, 16S sequencing of tissue samples demonstrated dominance by one or two genera that corresponded to the clinically isolated pathogens identified by airway cultures in 80% of samples (Figure S4). The high concordance with clinical isolates established that whenever there is high bacterial load, even basilar tissue samples can reliably capture the bacteria present, despite their smaller biomass compared to corresponding airway-based samples. Importantly, for the majority of IPF (and COPD) parenchymal samples, typical respiratory bacteria (commensal or pathogenic) were conspicuously absent (Figure S3).

### Bacterial load and composition in IPF tissue samples is associated with clinical outcomes

We noted a distinct subgroup of IPF samples with high bacterial load by qPCR, in the range of bacterial loads observed for CF samples. IPF samples within the highest bacterial load tertile had the lowest alpha diversity (Figure 4A) and were taxonomically distinct from samples from the other two tertiles (Figure 4B). Upon retrospective review of associated clinical variables, we found that patients receiving systemic antibiotics within the last three months had higher bacterial load compared to those not receiving antibiotics (p=0.04) (Figure 4C), possibly signaling a recent clinical deterioration that prompted an antibiotic prescription. Patients diagnosed with AE-IPF also had higher bacterial load (p=0.03) and were less likely to have survived to undergo lung transplantation (p=0.02) (Figure 4C).

**Figure 4.**
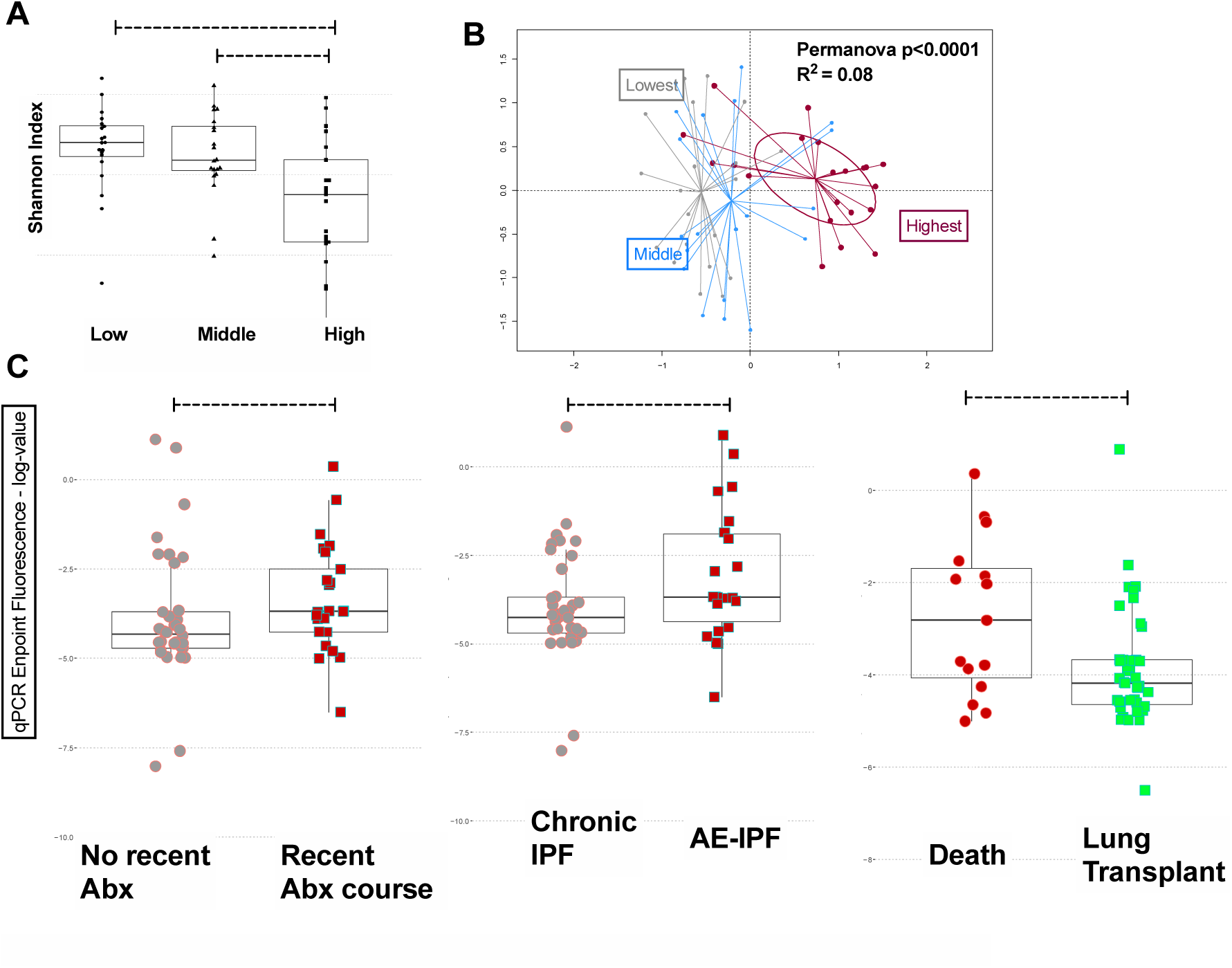
(A) Alpha diversity, as measured by Shannon index, of IPF basilar parenchymal tissue stratified by tertiles of bacterial load. (B) Beta diversity, as measured by Bray-Curtis dissimilarity, in IPF basilar parenchymal tissue stratified by tertiles of bacterial load. (C) Mean bacterial burden by qPCR endpoint fluorescence in basilar parenchyma IPF tissue stratified by clinical outcomes: by whether patients received a recent (within the preceding 90 days) antibiotic prescription, by whether patients had an AE-IPF at the time of lung explantation (transplant or death) and by whether patients died or received a lung transplant. P-values are obtained by Wilcoxon tests. *=p<0.05, ***=p<0.001, ****=p<0.0001. Abx, antibiotics; IPF, idiopathic pulmonary fibrosis; AE-IPF, acute exacerbation of IPF; MDS1, primary NMDS axis; MDS2, secondary NMDS axis; NMDS, non-metric multidimensional scaling.

We then agnostically examined for the presence of distinct microbial composition clusters in basilar IPF tissue samples by DMM. Two clusters offered the best model fit. Cluster 1 (71% of samples) consisted of communities with low bacterial load, high alpha diversity and abundance of bacteria that are not typical members of the respiratory microbiome (e.g. *Bradyrhizobium* and *Methylobacterium*), which likely represent experimental contamination (Figure 5). Cluster 2 (29%) demonstrated high abundance of typical members of the microbiome of respiratory tract (*Streptococcus, Veillonella* or *Prevotella* genera), and communities with higher bacterial load (p<0.001) and lower alpha diversity (p<0.05) compared to cluster 1 (Figure 5). Membership in cluster 2 was associated with higher odds ratio for diagnosis of AE-IPF (OR=3.3 [0.9-12.2], p=0.04) and recent antibiotic prescription (OR=6.6 [1.8-29.5], p=0.002) and lower odds ratio for survival to lung transplantation (OR=0.16 [0.04-0.66], p=0.007). We did not find any association between bacterial load or DMM clusters by *MUC5B* genotypes in the subset of IPF patients who underwent lung transplantation.

**Figure 5:**
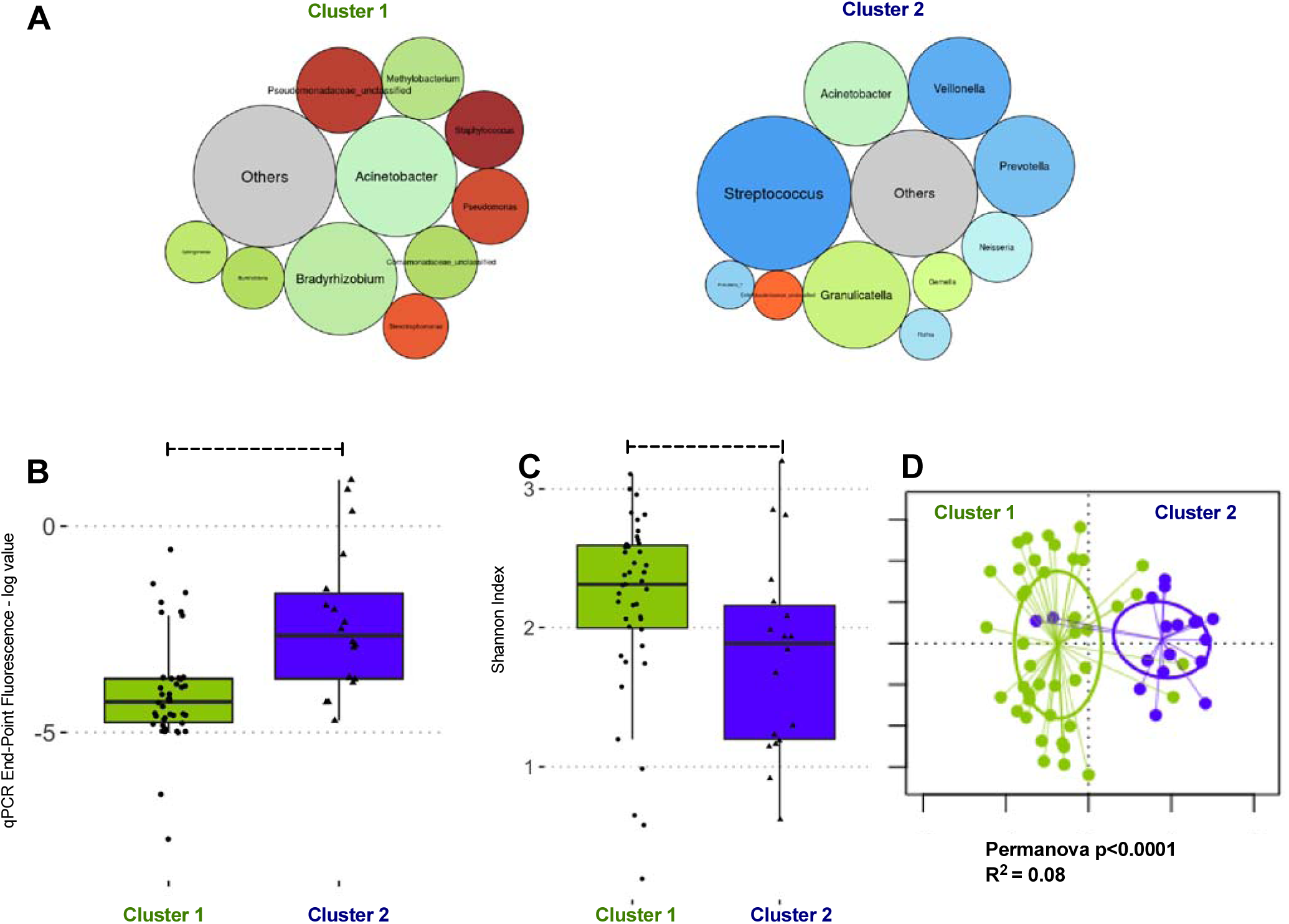
Dirichlet Multinomial Modeling clustering of IPF basilar tissue sample communities reveals two distinct taxonomic clusters. (A) Summary relative abundance for top 10 genera in each cluster, visualized as bubble plot (diameter of each circle corresponds to relative abundance of each taxon across all samples in the cluster). Cluster 1 (71% of samples) had high abundance for several genera that are not typical members of the respiratory microbiome and may represent procedural contamination (e.g. *Bradyrhizobium, Methylobacterium, Comamonadaceae*). Cluster 2 (29% of samples) had high abundance of typical members of the respiratory microbiome (*Streptococcus, Prevotella* and *Veillonella* genera). Genera beyond the top 10 genera demonstrated in these bubble plots were summarized to their overall relative abundance as a single bubble in grey and annotated “Others”. (B) Cluster 2 had significantly higher bacterial load compared to cluster 1 (log-transformed end-point fluorescence of 16S rRNA gene quantitative polymerase chain reaction). (C) Cluster 2 had lower alpha diversity (Shannon index) compared to cluster 1. (D) Principal coordinates analysis for visualization of beta-diversity differences (Bray-Curtis dissimilarity) between Cluster 1 and Cluster 2 samples. P-values are obtained by Wilcoxon tests. *=p<0.05; ***=p-value <0.001.

### Bacterial load and diversity do not differ along the apicobasal gradient of fibrosis

Examination of bacterial communities in IPF lungs along the typical apicobasal gradient of fibrosis seen in IPF revealed no difference in bacterial load by qPCR between apical and basilar tissue (Figure S5). Upon closer inspection of bacterial distribution, the majority of patient samples exhibited consistently low bacterial signals across all three lobes. Additionally, the taxonomic composition of communities was indistinguishable between each lobe (Permanova p-value for lobe non-significant). However, a distinct subgroup of six IPF lungs with high bacterial load (above 75^th^ percentile) was identified not only in the basilar but also in the middle/lingula and upper lobe samples (Figure S5). In these six cases, similar taxa were highly abundant in samples from all two or three lobes available, further underlying the notion for the presence of a patient-specific rather than a lobe-specific microbiome in IPF (Figure S6).

## DISCUSSION

In this study, we identified that basilar lung parenchyma samples have consistently decreased bacterial load compared to airway-based samples. End-stage parenchymal destruction as seen with advanced IPF and COPD represents a “microbiome desert” with the absence of identifiable respiratory bacterial communities in most cases. We also determined that while bacterial communities in IPF do not differ across the apicobasal gradient of fibrosis, a distinct subgroup of IPF patients demonstrated high bacterial load with abundance of typical respiratory microbiota, as well as worse clinical outcomes including acute exacerbations and death. Amongst the subgroup of IPF patients receiving a lung transplant, no association was detected between *MUC5B* genotypes and bacterial load, nor with DMM cluster assignment. These novel data generate further hypotheses regarding the potential of personalized microbiome-targeted therapies in IPF.

The spatial and temporal heterogeneity of UIP in IPF classically develop along an apicobasal gradient with fibrosis, with end-stage honeycombing appearing mostly in the subpleural lower lobes compared to the upper lobes.[16] Recent studies examining whether BAL-sampled microbiota vary among IPF patients with different radiographic features (e.g. honeycombing or traction bronchiectasis) did not reveal any significant associations.[9, 22] Similarly, we did not identify significant differences in the microbial communities of apical vs. basilar parenchymal samples, with the latter having greater extent of radiographic honeycombing. Instead, we found evidence of patient-specific, rather than lobe-specific microbiome in IPF. If local replication within the lower respiratory tract was a predominant source of origin for the lung microbiome, one might expect to see greater diversity across lobes. Our findings may indicate that microaspiration and dispersion of microbiota in the lower airways and parenchyma is the primary shaping force of microbial communities in IPF.

Our study utilized lung explants for excision of tissue specimens and acquisition of bronchial washings rather than obtaining trans-oral bronchoscopic samples as in prior investigations. Explanted tissue not only allowed us to examine the bacterial burden and communities in patients with advanced IPF that were unable to undergo research bronchoscopy, but also obviated the concern for bronchoscopic contamination by upper airway bacteria. Previous prospective analyses of IPF lung microbiota have focused on BAL within early-stage IPF patients, which may not be reflective of the microbiota present in those with the most advanced disease. Unlike previous studies,[7, 23] we did not identify an association between *Streptococcus* or *Staphylococcus* genus abundance and disease progression within our advanced IPF population. Instead, we utilized an unsupervised clustering approach to identify whether distinct meta-communities of lung microbiota existed in the IPF cohort. With agnostic DMM, we identified that 71% of samples had low bacterial burden and high alpha diversity, with several genera not typically associated with the respiratory tract, suggesting their possible origin from experimental contamination rather than true biologic presence. The remaining 29% of samples exhibited higher bacterial loads with typical respiratory microbiota abundance (*Streptococcus, Veillonella, Prevotella* genera). Notably, this cluster of patients with high loads of respiratory microbiota was associated with worse clinical outcomes, suggesting that high bacterial burden may impact clinical decline not only in early disease shown in previous studies,[7-9] but also in end-stage IPF.

Lung bacterial communities resemble those of the oral cavity in healthy individuals, though certain bacteria are significantly more abundant in the lung.[19] As oral samples were not collected from study patients, we were unable to ascertain the correlation between oral and lung microbiota in these patients. Given the high prevalence of gastroesophageal reflux disease (GERD) in IPF and the subsequent probable repeated micro-aspirations, higher correlation of lung and gut microbiome might be expected in IPF.[24, 25] While much of the association between GERD and IPF may be related to smoking, GERD has been implicated in many other respiratory diseases as well including asthma, COPD, bronchiolitis obliterans, and organizing pneumonia.[26-30] GERD was clinically present in 77% of our IPF patients (as defined by clinical history, esophagram and upper endoscopy results, and prescription for proton-pump inhibitor or H2 blocker), though the rate of silent micro-aspiration could be even higher. Indeed, increased micro-aspiration could account for the higher bacterial burden of BAL fluid in IPF compared to healthy controls previously identified. Further efforts to treat GERD, and more importantly prevent micro-aspiration events in IPF (rather than acid controlled focused therapy alone), may have a vital role in preventing the dysbiosis associated with worse clinical outcomes. However, our analyses by clinically-defined GERD did not reveal significant differences in microbial communities, similar to previous observations.[9]

Our study was limited by its retrospective design and utilization of a convenience dataset rather than a prospective cohort. All IPF samples are from patients with end-stage disease receiving care at a tertiary medical center, and thus may not reflect the general IPF population. Lung tissue remains a challenging biospecimen for microbiome work due to its low biomass relative to the amount of human DNA present, contamination risks, and readouts at levels near the detection limit of assays used. While we examined the bacterial composition of more anatomical regions compared to previous studies, the current lack of viral and fungal data limits our study from presenting a comprehensive representation of the microbiome in IPF. Noted associations with clinical outcomes, including the decreased likelihood of receiving a lung transplant and increased incidence of AE-IPF in patients with higher bacterial load, are hypothesis-generating only, given their retrospective nature. How to identify such patients while balancing the potential benefits of targeted antimicrobials with the risks of overtreatment and diagnostic procedures in patients with advanced lung disease remains unclear.

In summary, our analysis utilizes the distinct capacity of culture-independent sequencing techniques to discern that end-stage IPF lungs have limited biomass bacterial communities, without evidence of spatial heterogeneity across the apicobasal gradient of fibrosis. A subpopulation of patients with higher bacterial load and taxonomic species dominated by typical respiratory pathogens are more likely to die than undergo lung transplantation and may represent a crucial population towards whom microbiome-targeted interventions ought to be considered. The ongoing development of rapid, culture-independent methods for profiling microbiota holds the promise for personalized medicine approaches in IPF.

## Data Availability

All de-identified sequencing data have been submitted to Sequence Read Archive (SRA) database, with BioSample accession number of SAMN13906474-13906711.

## Acknowledgements

The authors would like to acknowledge The Center for Organ Recovery & Education (CORE) as well as organ donors and their families for the generous donation of tissue used in this study.

## Author contributions

Conception and design: GDK, MR, AM, BJM

Acquisition, analysis or interpretation of data: EV, HY, JCS, LY, SW, RN, DJK, SQ, XW, MMM, BM, AF, JKA, PVB, BJM, MR, AM, GDK

Clinical cohort characterization: GDK, EV, SW, MMM, DJK

Drafting of work and/or revising for important intellectual content: EV, HY, JCS, LY, SW, RN, DJK, SQ, XW, MMM, BM, AF, JKA, PVB, BJM, MR, AM, GDK

Final approval of version to be published; agreement to be accountable for all aspects of the work in ensuring that questions related to the accuracy or integrity of any part of the work are appropriately investigated and resolved: EV, HY, JCS, LY, SW, RN, DJK, SQ, XW, MMM, BM, AF, JKA, PVB, BJM, MR, AM, GDK

## Data sharing

All de-identified sequencing data have been submitted to Sequence Read Archive (SRA) database, with BioSample accession numbers of SAMN13906474-13906711.

